# Comprehensive investigation of DNA damage repair genes in children with cancer identifies *SMARCAL1* as novel osteosarcoma predisposition gene

**DOI:** 10.1101/2025.05.12.25325832

**Authors:** Ninad Oak, Wenan Chen, Alise Blake, Lynn Harrison, Martha O’Brien, Christopher Previti, Gnanaprakash Balasubramanian, Kendra Maass, Steffen Hirsch, Judith Penkert, Barbara C Jones, Kathrin Schramm, Michaela Nathrath, Kristian W Pajtler, David T.W. Jones, Olaf Witt, Uta Dirksen, Jiaming Li, Yadav Sapkota, Kirsten K Ness, Lillian M Guenther, Stefan M Pfister, Christian Kratz, Zhaoming Wang, Greg T Armstrong, Melissa M Hudson, Gang Wu, Robert J Autry, Kim E Nichols, Richa Sharma

## Abstract

**Background:** Recent large-scale genomic sequencing studies reveal that 5-18% of children with cancer harbor pathogenic variants (PV) in known cancer predisposing genes (CPG). However, DNA damage repair (DDR) genes, which are frequently somatically altered in pediatric tumors, have not been systematically examined as a source of novel cancer predisposing signals.

**Methods:** To address this gap, we interrogated 189 genes across six DDR pathways for the presence of PV among 5,993 childhood cancer cases and 14,477 adult non-cancer controls. PV were defined as rare (allele frequency <0.05% in the gnomAD v2.1 non-cancer subset), nonsense, frameshift, affecting canonical splice sites, and missense with REVEL score >0.7. Using logistic and firth regression, we identified genes with statistically enriched PV and replicated findings among 1,494 additional childhood cancer cases across three independent cohorts.

**Findings:** Analysis across all cancers revealed enrichment of *TP53* PV (0.6%, false discovery rate [FDR]_logistic_=0.0066, FDR_Firth_=0.0064). Cancer-specific analyses confirmed previously identified associations for germline *TP53* PV in adrenocortical carcinoma (37%, FDR_logistic_<0.0001, FDR_Firth_=0) and high-grade glioma (2.4%, FDR_logistic_=0.0022, FDR_Firth_=0.1082), as well as *BARD1* PV in neuroblastoma (1.2%, FDR_logistic_=0.0341, FDR_Firth_=0.2682). Three novel gene-tumor associations were identified, including *POLL* PV in Ewing sarcoma (1.7%, FDR_logistic_=0.0319, FDR_Firth_=0.3101), *SMC5* PV in medulloblastoma (1.6%, FDR_logistic_=0.0005, FDR_Firth_=0.0499) and *SMARCAL1* PV in osteosarcoma (2.6%, FDR_logistic_=0.0250, FDR_Firth_=0.2180). Among these putative CPG, *SMARCAL1* PV were enriched in osteosarcoma across each of the replication pediatric cancer cohorts (2.5%, P_Fisher_ <0.0001). All three osteosarcomas with available tumor data exhibited deletion of the wild-type *SMARCAL1* allele.

**Interpretation:** Our study identifies *SMARCAL1* PV as a predisposing factor for osteosarcoma, providing insights into tumor biology and creating opportunities for development of novel therapeutic, surveillance, and preventive interventions for this aggressive childhood cancer.

## Background

As the initiating genetic events in tumorigenesis, germline variants in cancer predisposing genes (CPG) perturb cell growth and differentiation to set the stage for malignant transformation. Accordingly, the study of CPG and associated hereditary syndromes provides critical insights into normal physiology and cancer biology. To this end, recent sequencing studies have revealed that up to 18% of children with cancer harbor an underlying genetic predisposition.^1–4^ However, 40–80% of children with cancer have family histories and/or clinical features concerning for cancer predisposition but lack a causal genetic diagnosis.^1,5^ This observation suggests that additional CPG remain to be discovered and their association with cancer phenotypes further elucidated.

Prior investigations have primarily included children with specific tumor types (e.g., high risk solid or central nervous system [CNS] tumors, relapsed cancers) and examined for pathogenic variants (PV) in known CPG. Nevertheless, expanding the scope of germline analyses to include children with a broader array of cancers and additional cancer-associated genes is crucial to identify the missing heritable factors underlying childhood tumor formation. The identification of novel CPG and predisposing variants is also central to improving the outcomes for affected children as it enables development of targeted cancer therapies, genetic counseling and testing of relatives, and improves approaches to cancer surveillance and risk reduction.^6,7^

Somatic alterations impacting DNA damage repair (DDR) genes are prominent drivers of high grade pediatric tumors.^2^ In addition, germline PV impacting selected DDR genes have been linked to several highly penetrant childhood cancer predisposition syndromes (CPS), including Li-Fraumeni syndrome, ataxia telangiectasia, Fanconi anemia, and replication repair deficiency.^2,8^ Germline PV in DDR pathway genes have also been implicated in development of subsequent malignant neoplasms in long-term survivors of childhood cancer, especially those previously exposed to higher doses of ionizing radiation, anthracyclines or alkylating agents.^9^ Despite these prior observations and to the best of our knowledge, an unbiased assessment of DDR pathway genes and their role in development of primary cancers in children has not been conducted.

To this end, we generated a harmonized dataset of germline variants from 5,993 childhood cancer cases and 14,477 adult non-cancer controls. We then conducted rare variant gene burden analysis using a curated set of 189 DDR genes with the aim to identify novel CPG that could account for the missing heritability of childhood cancer. Novel gene-cancer associations were replicated using three independent pediatric cancer cohorts and available tumor data.

## Methods

### Patient Cohorts

The discovery cohort consisted of 5,993 children with cancer across five large scale sequencing studies, including the Pediatric Cancer Genome Project (PCGP, phs000409),^10^ National Cancer Institute Therapeutically Applicable Research to Generate Effective Treatments initiative (NCI-TARGET, phs000218),^11^ St. Jude Lifetime Cohort Study (SJLIFE),^12^ Genomes for Kids (G4K),^3^ and St. Jude Real-Time Clinical Genomics study (RTCG) (**Figure 1A**, **Supplementary Table S1**). The control cohort for discovery comprised 14,477 adults without cancer from the 1000 Genomes Project^13^ and Alzheimer’s Disease Sequencing Project (phs000572).^14^ This study was approved by the Institutional Review Board at St. Jude Children’s Research Hospital (No. 20-0379). For replication of novel CPG, we queried three independent pediatric cancer cohorts, including the Childhood Cancer Survivorship Study (CCSS, phs001327),^15^ INdividualized Therapy FOr Relapsed Malignancies in Childhood (INFORM)^16^ and the German Childhood Cancer Registry (GCCR). The control cohort for replication included adults without cancer from gnomAD v2.1^17^.

**Figure 1.**
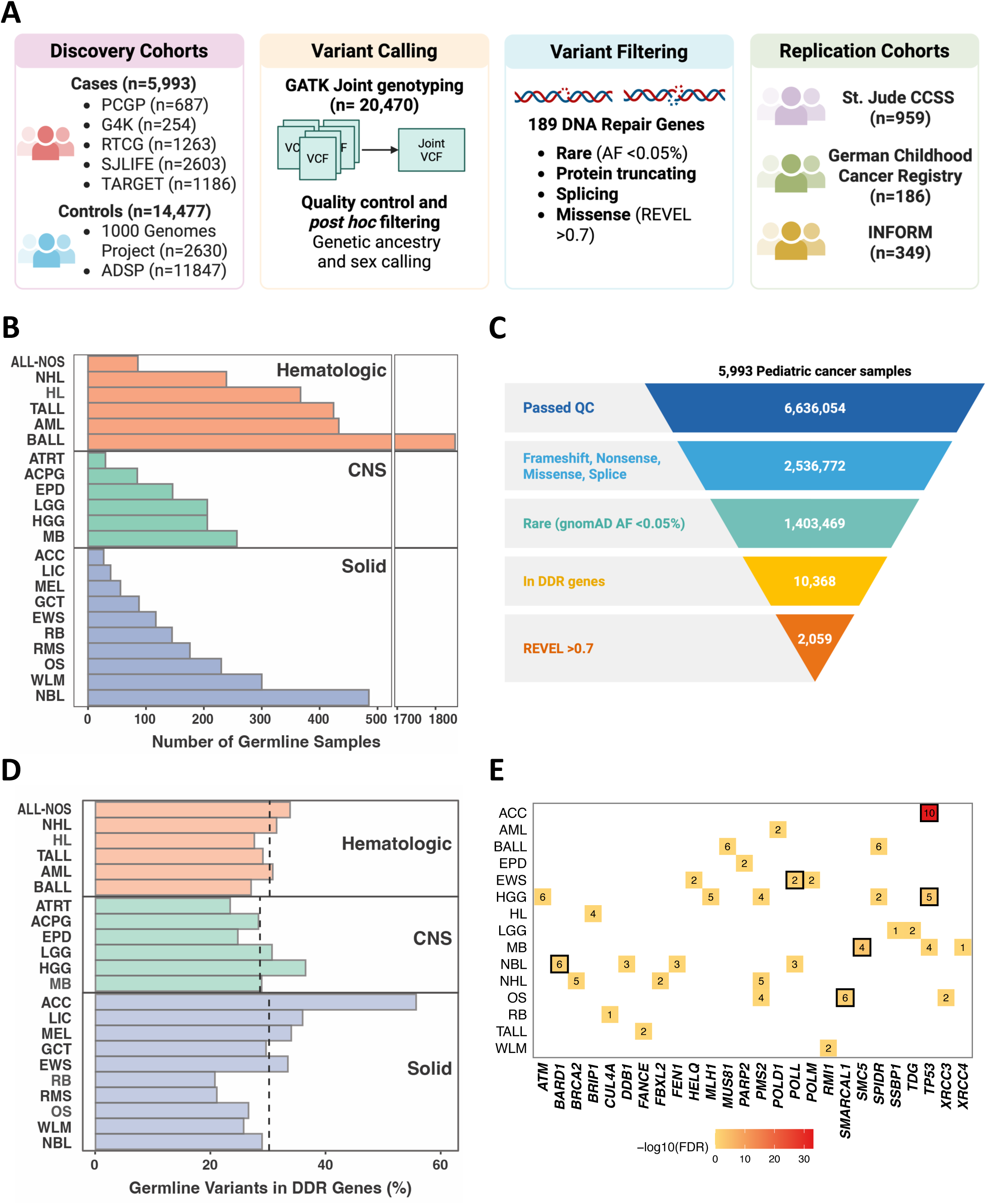
Germline DDR gene variants across pediatric cancers. **(A)** Study design and workflow. The discovery cohort comprised 5,993 pediatric cancer patients from the PCGP, G4K, RTCG, SJLIFE, and TARGET cohorts, with 14,477 controls from the 1000 Genomes Project and Alzheimer’s Disease Sequencing Project (ADSP). Germline variants were called using GATK joint genotyping (n=20,470) and underwent quality control and post hoc filtering along with genetic ancestry and sex determination. Variant filtering focused on 189 DNA repair genes, selecting rare variants (allele frequency <0.05%) predicted to be deleterious, including protein-truncating, splicing, and high-confidence missense variants (REVEL >0.7). Replication was performed using independent cohorts, including CCSS, GCCR, and INFORM. **(B)** Number of jointly called germline whole-exome sequencing samples across 22 pediatric cancers in the discovery cohort. **(C)** Variant filtering pipeline. Among 5,993 pediatric cancer samples, 6,636,054 variants passed quality control. Filtering by predicted functional impact (frameshift, nonsense, missense, and splicing) yielded 2,536,772 variants, of which 1,403,469 were rare (gnomAD v2.1 non-cancer popmax AF <0.05%). Restriction to DNA damage repair (DDR) genes identified 10,368 variants, with 2,059 meeting the pathogenicity threshold (protein truncating, splicing, or damaging missense-REVEL >0.7). **(D)** Prevalence of germline DDR gene variants across cases with 22 pediatric cancer types is shown. Median prevalence across the three major cancer classes (hematologic: red, CNS: green, solid: blue) is shown with a dashed line. **(E)** Gene-level burden of pathogenic germline variants. All associations with FDR <0.25 are shown, whereas those with FDR <0.05 are highlighted with a black border. The color intensity represents -log10[FDR]). The number of variants for respective gene-cancer pairs is indicated. Abbreviations: ACC= Adrenocortical Carcinoma; ACPG= Adamantinomatous Type Craniopharyngioma; ALL-NOS= Acute Lymphoblastic Leukemia-Not Otherwise Specified; AML= Acute Myeloid Leukemia; ATRT= Atypical Teratoid/Rhabdoid Tumor; BALL= B-Cell Acute Lymphoblastic Leukemia; CCSS= Childhood Cancer Survivorship Study; EPD= Ependymoma; EWS= Ewing Sarcoma; FDR = false discovery rate; G4K= Genomes4Kids; GCCR= German Childhood Cancer Registry; GCT= Germ Cell Tumor; HGG= High-Grade Glioma; HL= Hodgkin Lymphoma; INFORM= INdividualized Therapy FOr Relapsed Malignancies in Childhood; LGG= Low-Grade Glioma; LIC= Liver Cancer; MB= Medulloblastoma; MEL= Melanoma; NBL= Neuroblastoma; NHL= Non-Hodgkin Lymphoma; OS= Osteosarcoma; PCGP = Pediatric Cancer Genome Project; RB= Retinoblastoma; RMS= Rhabdomyosarcoma; RTCG= (St.Jude’s) Real-time Clinical Genomics; SJLIFE= St. Jude Lifetime Cohort; TALL= T-Cell Acute Lymphoblastic Leukemia; WLM= Wilms Tumor.

### Variant Calling and Filtering for the Discovery cohort

Germline whole exome sequencing (WES) reads from cases and controls were mapped to GRCh37 with Burrow-Wheeler Aligner followed by joint variant calling using GATK best practices workflow with modifications (**Supplementary Methods**). Downstream analyses were restricted to germline variants in 189 genes that function in DDR (**Supplementary Table S2**).^8,9^ We filtered to retain rare damaging (i.e., pathogenic) variants (PV) if the they fulfilled the following criteria: 1) minor allele frequency (AF) <0.05% in gnomAD v2.1 (non-cancer subset);^17^ 2) nonsense or frameshift variants; 3) missense variants with REVEL scores >0.7; and 4) canonical splice site variants. Matched tumor data for specific samples in our discovery cohort were analyzed as described in the **Supplementary Methods**.

### Statistical Analysis

We performed a gene-based burden analysis for rare damaging germline variants in DDR genes in the discovery pediatric cancer cohort versus non-cancer controls. Twenty-two cancer types with at least 25 cases were considered for this analysis. For gene burden analysis, we applied logistic regression (glm-logit, R package stats v3.6.2; hereafter represented as false discovery rate [FDR]_logistic_) and Firth regression (R package logistf v1.26; represented as FDR_Firth_ with odds ratio [OR]) using the first five principal components as co-variates to account for genetic ancestry. We computed nominal p-values using the Chi-squared and Walds test, followed by correction for multiple comparisons using the Benjamini & Hochberg method. Significance was determined as FDR <0.05. For replication of statistically significant gene-cancer associations, we queried relevant pediatric cancer cases in CCSS, INFORM, GCCR, and adult non-cancer controls (gnomAD v2.1) followed by filtering germline PV based on criteria similar to the discovery cohort. Fisher exact test was used to evaluate for enrichment of damaging germline variants in respective cancer types. Comparison of age at cancer diagnosis between DDR germline variant carriers and non-carriers was completed as described in the **Supplementary Methods**.

## Results

### Germline variants in DDR genes across tumor types

The discovery cohort included 5,993 children and adolescents with 22 cancer subtypes, broadly classified as hematologic, solid, and CNS tumors (**Figure 1A, B**). The median age at cancer diagnosis was 6 years (4 days to 32 years) with most cases (67%) of European ancestry (**Supplementary Table S3**). Among the 6,636,054 germline variants filtered from cases, 2,059 rare, protein truncating, splicing, or predicted damaging missense variants among 189 DDR genes were retained (**Figure 1C, Supplementary Table S4**). The prevalence of putative PV among the 189 DDR genes in cases was comparable to controls (26.7%; p=0.07; **not shown**). Additionally, the frequency of PV was similar across hematologic (mean prevalence 29.9% [range 26.9% - 33.7%]), solid (31.1% [20.7% - 55.6%]), and CNS tumors (28.7% [23.3% - 36.4%]; p=0.8, Kruskal-Wallis test) but varied across cancer subtypes, with the lowest prevalence in retinoblastoma (20.7%) and highest in adrenocortical carcinoma (ACC, 55.6%) (**Figure 1D**).

### Significant germline associations in DDR genes

Overall, 1,697 of 5,993 (28.3%) childhood cancer cases harbored putative PV in one or more DDR genes. Analyses across all cancers revealed significant enrichment of PV in *TP53* compared to jointly-called adult non-cancer controls (n=14,477) (0.6%, FDR_logistic_=0.0066; FDR_Firth_=0.0064, OR=2.8 [95% CI: 1.7 - 4.6]), supporting its critical role in maintaining genome stability (**Supplementary Table S5)**. We next tested for enrichment of damaging germline variants in DDR genes across the 22 cancer subtypes in our discovery cohort. We observed enrichment of *TP53* PV in ten of 27 ACC cases (37%, FDR_logistic_<0.0001; FDR_Firth_=0, OR=289.2 [120.7 - 673.5]) and five of 206 high-grade glioma (HGG) cases (2.4%, FDR_logistic_=0.0022; FDR_Firth_=0.1082, OR=11.1 [3.9 - 26.0]) (**Figure 1E**, **Table 1**, **Supplementary Table S5, S6**). Both germline *TP53*-tumor associations have previously been described and serve as internal validation of our analytic pipeline.^1,18^ Of note, the founder variant, *TP53*:p.R337H, common in southern Brazil, did not meet our filtering criteria (REVEL=0.693) and therefore, was excluded from this analysis. Among the 14 unique germline PV in *TP53*, six were protein truncating or splicing variants and the remaining were missense variants reported to negatively impact transcriptional activity (**Figure 2A**).^19,20^ Tumor WES or WGS data were available for each of the germline *TP53*-mutated ACC and HGG, which showed a somatic second hit affecting the remaining *TP53* allele in all cases (**Supplementary Figure S1A**).

**Figure 2:**
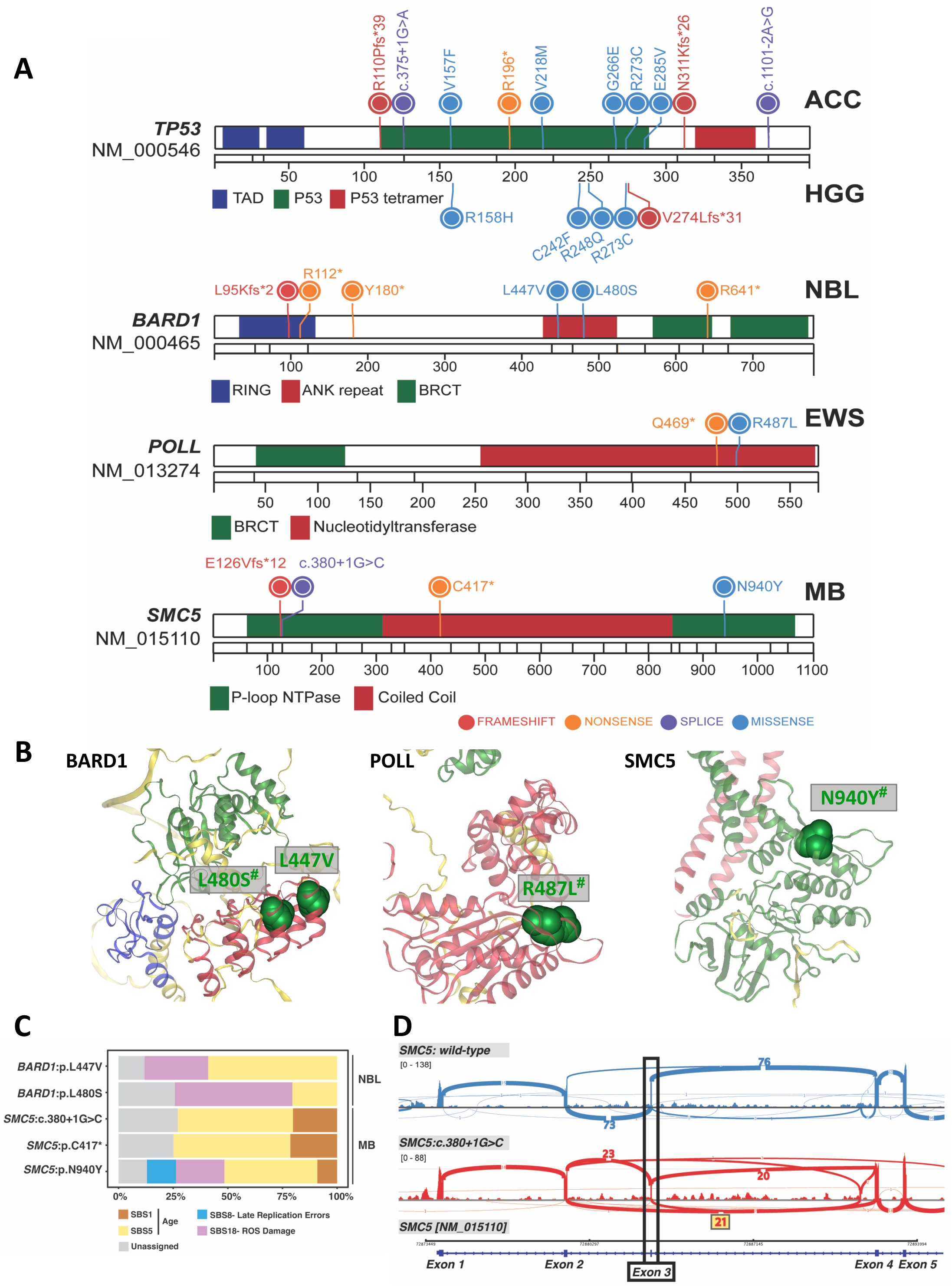
Germline mutations in DDR genes and their putative functional impacts. **(A)** Distribution of predisposing variants along the protein sequences of *TP53*, *BARD1*, *POLL*, and *SMC5* across adrenocortical carcinoma (ACC), high-grade glioma (HGG), neuroblastoma (NBL), Ewing sarcoma (EWS), and medulloblastoma (MB), respectively. Variants are categorized by type: frameshift (red), nonsense (orange), splice-site (purple), and missense (blue). **(B)** Structural mapping of selected missense variants in *BARD1*, *POLL*, and *SMC5* as predicted by AlphaFold. Germline variants such as p.L480S and p.L557V in the ankyrin repeat domain of BARD1, p.R487L in nucleotidyltransferase domain of POLL and p.N940Y in P-loop NTPase domain of SMC5 are located in regions critical for protein stability and/or function. ^#^ denotes variants predicted as P/LP by AlphaMissense. **(C)** Somatic DNA mutational signature analysis across five cases with whole genome sequencing data was performed using signature.tools.db. Bar plots display the proportion of COSMIC (v3) single-base substitution (SBS) mutational signatures, including SBS1 and SBS5 (aging-related), SBS8 (late replication errors), and SBS18 (reactive oxygen species damage). Unassigned mutational signatures are also shown. **(D)** Tumor RNAseq from the medulloblastoma (MB) case with the germline *SMC5*:c.380+1G>C splicing variant. The top panel (blue) represents normal splicing in another MB case without an *SMC5* alteration, while the bottom panel (red) shows aberrant splicing in the *SMC5*:c.380+1G>C germline variant positive case. The variant leads to skipping of exon 3, as indicated by disrupted exon-exon junctions and altered splicing events, suggesting a pathogenic impact of the mutation.

**Table 1.**
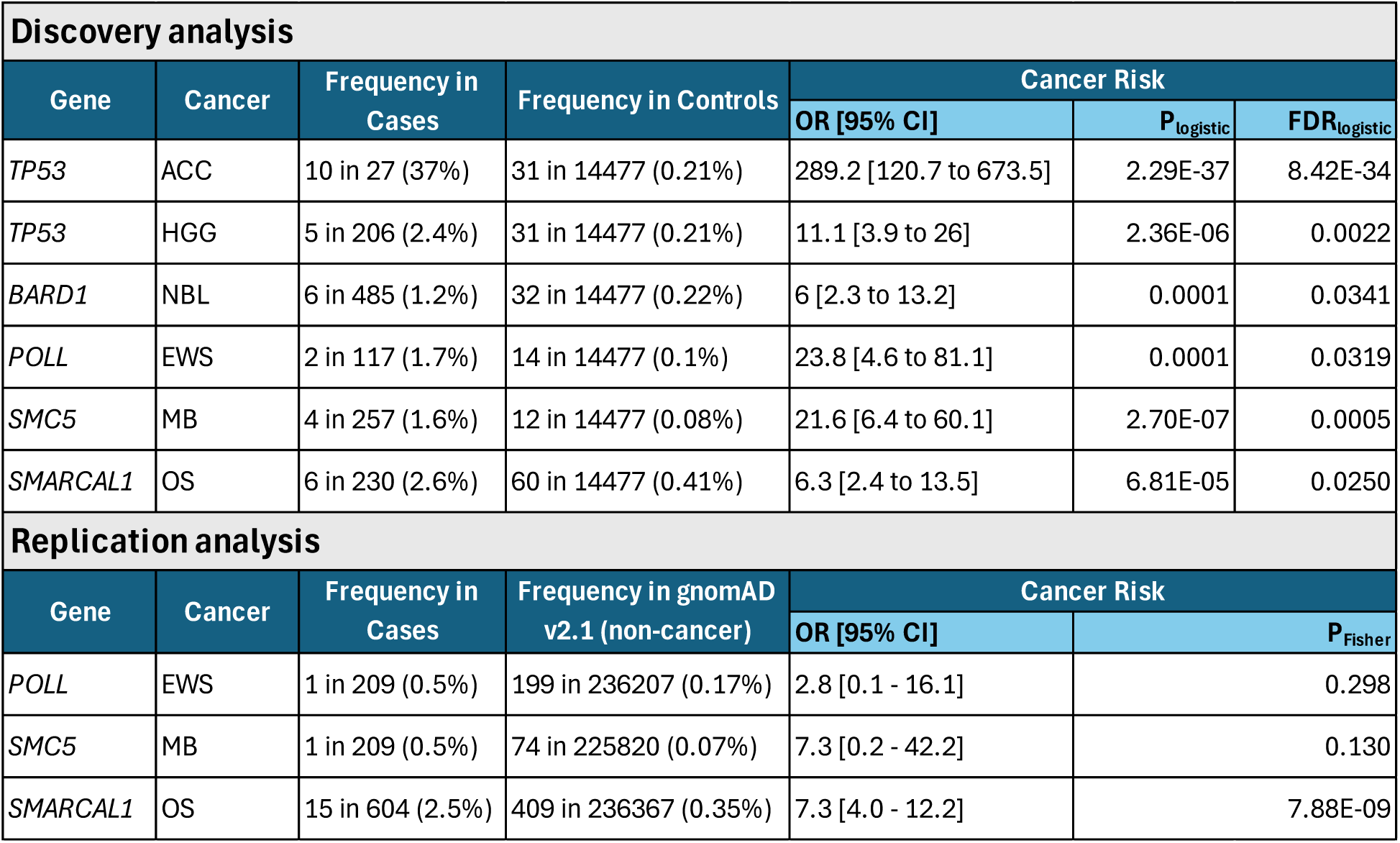
Significant cancer-gene associations from the discovery and replication cohorts.

The *BARD1* gene, which encodes BRCA1-associated RING domain 1, has emerged as a neuroblastoma (NBL) predisposition gene.^21^ Similar to prior reports, we observed germline *BARD1* variants in six of 485 NBL cases (1.2%; FDR_logistic_=0.0341; FDR_Firth_=0.2682, OR=6.0 [2.3 - 13.2]), including four predicted as truncating and two that were missense (**Figure 2A**, **Supplementary Tables S5, S6**). The missense variants (p.L480S, p.L447V) are located in the ankyrin repeat domain of BARD1, a region in which mutations cause HR deficiency (**Figure 2B**).^22^ Tumor WES data were available for all six NBL cases and WGS data for two cases. Second somatic hits affecting *BARD1* were not observed in any of these cases (**Supplementary Figure S1A**). In the two tumors with WGS data, DNA mutation signature analysis revealed presence of SBS18, associated with reactive oxygen species damage, a signature observed in NBL (**Figure 2C**).^23^ RNAseq data were available for three tumors, one of which (germline p.Y180*) showed *BARD1* expression at less than 10^th^ percentile compared to non-*BARD1* mutated NBL tumors (**Supplementary Figure S1B**). While this finding suggests loss-of-heterozygosity (LOH) in the tumor, tumor WGS data were not available to corroborate this possibility.

In addition to verifying known associations, we identified three novel gene-cancer associations, including *POLL* in Ewing sarcoma (EWS), *SMC5* in medulloblastoma (MB), and *SMARCAL1* in osteosarcoma (OS) (**Table 1**, **Supplementary Table S5**). Two of 117 (1.7%; FDR_logistic_=0.0319; FDR_Firth_=0.3101, OR=23.8 [4.6 - 81.1]) EWS cases exhibited germline damaging variants in *POLL* (**Figure 2A, Supplementary Table S6**). *POLL* encodes the DNA polymerase lambda, which performs 3’-end extension during non-homologous end joining (NHEJ) and base excision repair (BER). Both p.Q469* and p.R487L are located in the nucleotidyltransferase domain within the DNA binding groove of POLL that is important for gap filling during NHEJ (**Figure 2B)**. Neither variant is included in ClinVar and the missense variant p.R487L is predicted as LP by AlphaMissense. Tumor WES and RNAseq data were available for the p.Q469*-associated tumor, which revealed the EWS somatic driver, *EWSR1:FLI1*, and no additional SNVs in *POLL* (**Supplementary Figure S1A, Supplementary Table S6**), which is consistent with heterozygosity in the tumor and supported by the observed 63^rd^ percentile *POLL* expression noted by tumor RNAseq (**Supplementary Figure S1B**).

Four of 257 MB cases (1.6%; FDR_logistic_=0.0005; FDR_Firth_=0.0499, OR=21.6 [6.4 - 60.1]) exhibited germline PV in *SMC5*, the gene encoding Structural Maintenance of Chromosome 5 (**Figure 2A**). SMC5 is important for DNA replication, repair and chromosome maintenance with biallelic germline alterations associated with the neurodevelopmental disorder, Atelis syndrome-2.^24^ Interestingly, all four germline *SMC5* mutant cases were of the group 3 MB subtype. The association of germline *SMC5* PV with MB was even stronger when analyses were restricted to include only group 3/4 MB (four of 88 cases, 4.5%; FDR_logistic_<0.0001; FDR_Firth_=0.0011, OR=54.6 [16.0 - 156.0]) (**Supplementary Table S5**). The p.E126Vfs*12 and c.380+1G>C, located in the N-terminus P-loop NTPase domain, and the p.C417* variant, located in the coiled-coil domain, are predicted to result in nonsense-mediated decay (NMD). The missense variant p.N940Y was located in the C-terminal P-loop NTPase domain of SMC5, important for DNA binding and ATP-driven loop extrusion by the SMC5/6 complex (**Figure 2A-B**). Although none of the *SMC5* variants were reported in ClinVar, p.N940Y is predicted to be LP by AlphaMissense. Tumor RNAseq and WES data were available for all four *SMC5* germline variant carriers, whereas WGS was available for three. Tumor *SMC5* expression was variable across the four cases. The MB tumor associated with germline c.380+1G>C, resulting in an out-of-frame transcript, showed exon 3 skipping in ∼50% of *SMC5* transcripts (**Figure 2C**) and *SMC5* expression at <10^th^ percentile compared to *SMC5* wild-type MB (**Supplementary Figure S1B**). Due to lack of tumor WGS data for this case, we could not determine whether there was a deletion affecting the remaining *SMC5* allele. We identified a second somatic hit, *SMC5*:p.Q357K, in the p.N940Y germline-mutated MB but could not establish *cis* versus *trans* allelic configuration of the two variants. Of note, this tumor exhibited SBS8, a DNA mutational signature associated with late replication errors in cancer (**Figure 2C**), supporting SMC5’s role in mitotic progression.^25^

Finally, we identified six of 230 OS cases (2.6%; FDR_logistic_=0.0250; FDR_Firth_=0.2180, OR=6.26 [2.4 - 13.5]) with germline *SMARCAL1* variants (**Figure 3A, Supplementary Tables S5, S6**). *SMARCAL1* encodes the SNF2 related Chromatin Remodeling Annealing Helicase, which resolves stalled replication forks and secondary DNA structures to support DNA replication and repair.^26^ We identified four protein-truncating variants (p.R114Qfs*4, p.L139Efs*3, p.L397Rfs*40, p.Q653*) that are predicted to undergo NMD and two missense variants (p.R820H, p.R490C) that are located in the SMARCAL1 helicase domain. The p.R820H variant was classified as P by ClinVar and predicted as LP by AlphaMissense, while p.R490C was classified as a VUS in ClinVar and ambiguous by AlphaMissense (**Figure 3B-C, Supplementary Table S6**). Considering that OS is commonly observed in Li-Fraumeni syndrome, we queried and confirmed the absence of germline *TP53* mutations in the six germline *SMARCAL1*-mutated cases. Further, none of the OS cases harbored germline variants in a list of 60 known CPG associated with autosomal dominant CPS of moderate to high penetrance.^27^ Matched tumor WES and RNAseq data were available for two cases, which did not reveal a somatic second hit (SNV) in *SMARCAL1*. The germline p.R114Qfs*4-mutated OS harbored a somatic *ATRX*:p.P717Hfs*4 mutation. Tumor RNAseq from this case showed unaffected *SMARCAL1* expression, while the germline p.Q653*-mutated case demonstrated reduced *SMARCAL1* RNA expression <1^st^ percentile. However, additional genomic alterations associated with this reduced expression could not be ascertained in the absence of tumor WGS data (**Figure 3B**, **Supplementary Table S6**, **Supplementary Figure S1B)**.

**Figure 3:**
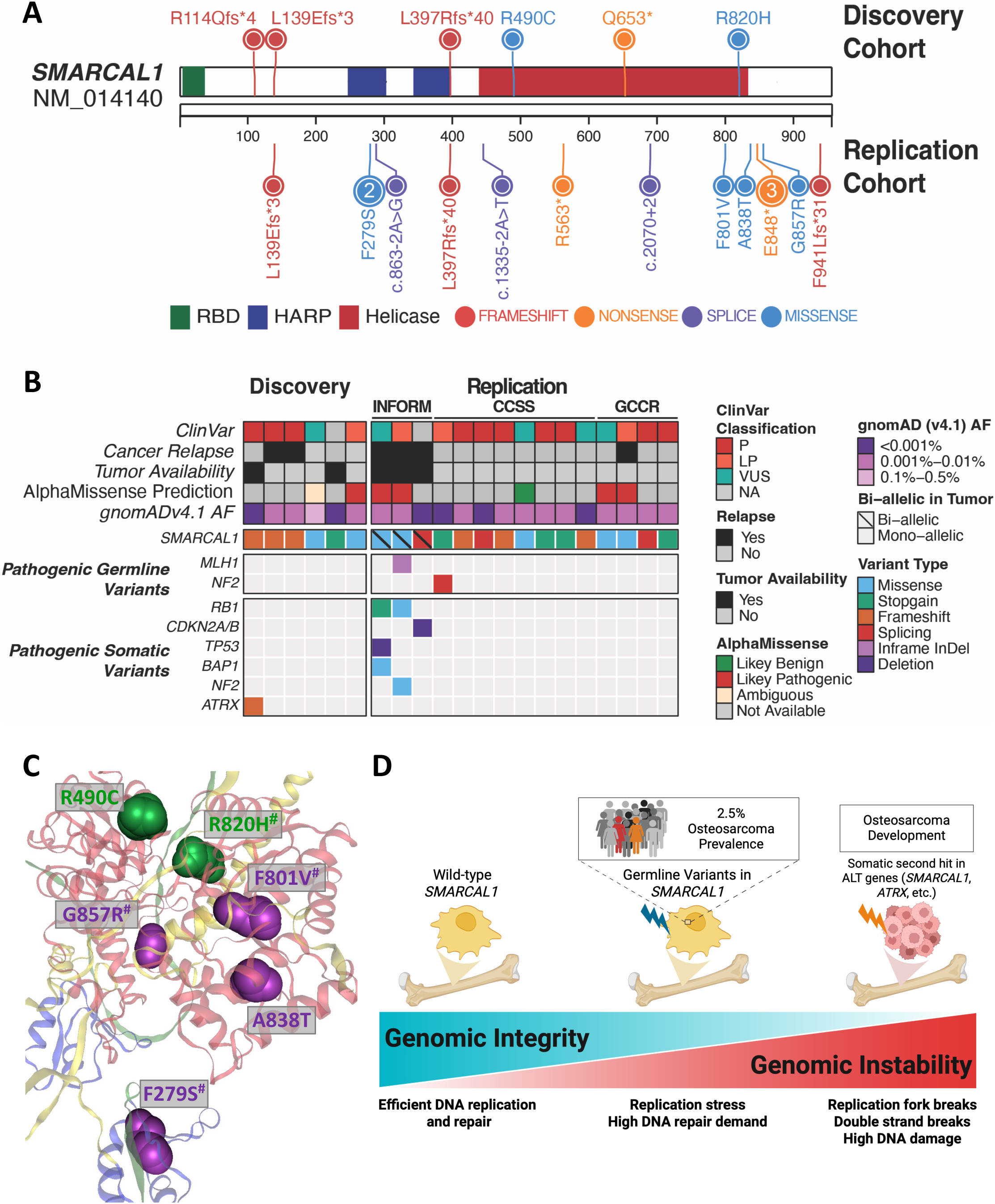
Characterization of germline *SMARCAL1* variant-positive osteosarcoma (OS). **(A)** Schematic representation of SMARCAL1 (NM_014140) protein with predisposing variants from the discovery cohorts (top) and replication cohorts (bottom). Protein domains RBD-RPA binding domain (green), HARP-HepA-related protein (blue) and Helicase (red) are shown. Variants are categorized by mutation type with frameshift (red), nonsense (orange), splice-site (purple), and missense variants (blue). **(B)** Heatmap representation of *SMARCAL1* variants in discovery and replication cohorts. Data include pathogenicity predictions based on ClinVar classification, REVEL scores, AlphaMissense predictions, and gnomADv2.1 allele frequencies. Additional pathogenic germline variants in SJCPG_60_ genes or somatic driver events are shown along with annotations for biallelic status (\), tumor relapse status and tumor availability. **(C)** SMARCAL1 protein structure predicted by AlphaFold. Missense variants in discovery (green) and replication (purple) cohorts of OS are shown. Protein domains HARP-HepA-related protein (blue) and Helicase (red) are shown. ^#^ Denotes variants predicted as LP by AlphaMissense. **(D)** Proposed model of SMARCAL1-mediated OS predisposition. SMARCAL1 is important for accurate DNA replication and repair to reinforce genome integrity. Approximately, 2.5% of osteosarcoma cases carry a predisposing variant in *SMARCAL1*, negatively impacting SMARCAL1 function and exacerbating genome instability with tumor acquisition of somatic second hits in genes known to permit ALT (*SMARCAL1* LOH, *ATRX* inactivation), resulting in OS development.

### Clinical features associated with germline DDR gene variants identified in the Discovery cohort

Patients with germline CPG PV are often younger at tumor onset than individuals with sporadic cancers. Therefore, we analyzed the ages of cancer onset in carriers versus non-carriers of damaging DDR gene variants for the significant gene-cancer associations. Through this analysis, we observed a younger age of onset of 20 months in ACC cases carrying *TP53* PV versus 60 months (Wilcoxon rank-sum test p<0.05) in non-carriers (**Supplementary Table S7**). We did not find significant differences in ages of cancer onset for any of the other significant gene-cancer associations, or any DDR PV carriers across all cancers when compared to non-carriers. Family history information was available for 17 of 33 (52%) patients with significant gene-cancer associations, eight of whom had a positive family history of cancer, defined as having ≥1 first- or second-degree relatives under 50 years of age with cancer or tumor (excluding cervical and non-melanoma skin cancer; **Supplementary Table S6**). However, none of the relatives from cases with a positive family cancer history had tumors similar to the cases described here.

### Replication of *SMARCAL1* as a novel osteosarcoma predisposition gene

To replicate our novel gene-tumor associations, we queried three additional pediatric cancer cohorts (CCSS, GCCR, and INFORM). *POLL* did not replicate and *SMC5* reached significance in an inadequately powered GCCR MB cohort (one of 30; 3.3%; P_Fisher_=0.020, OR= 52 [1.3 – 309.1]. Importantly, we confirmed significant enrichment of damaging *SMARCAL1* variants in 15 children with OS across CCSS (eight of 272 cases; 2.9%; Fisher exact P_Fisher_<0.0001, OR= 8.6 [3.7 - 17.3]), GCCR (four of 135 cases; 3%; P_Fisher_=0.001, OR= 8.7 [2.3 - 22.6]), and INFORM (three of 197 cases; 1.5%; P_Fisher_ =0.032, OR= 4.4 [0.9 - 13.1]) compared to 0.35% (409 in 118,184) in adult non-cancer controls (gnomAD v2.1) (**Supplementary Table S8**). These 15 cases harbored 12 unique germline variants, two of which were also observed in the discovery cohort (p.L139Efs*3, p.L397Rfs*40). Among the 10 remaining *SMARCAL1* variants, three were protein-truncating (p.F941Lfs*31, p.R563*, p.E848*), three canonical splice-site (c.863-2A>G, c.1335-2A>T, c.2070+2dup) and four missense (p.F801V, p.A838T, p.G857R, p.F279S). Three of the missense variants (p.F801V, p.A838T, p.G857R) are clustered within the SMARCAL1 helicase domain that is critical for DNA binding, while one (p.F279S) is located in the Hep-A-related protein (HARP) domain, essential for replication fork stabilization (**Figure 3C**).^26^ Five of these 10 variants are reported as P or LP and four as VUS in ClinVar, while one is unreported (**Supplementary Table S9)**. Three missense variants (p.F801V, p.G857R, p.F279S) are predicted to be LP by AlphaMissense, suggesting an adverse impact on SMARCAL1 structure (**Figure 3C**, **Supplementary Table S9)**. None of the 15 *SMARCAL1* germline variant carriers from these replication cohorts harbored a PV in *TP53*; however, two cases harbored PV affecting other CPG: one in *NF2* (c.243-2A>C, CCSS) and one in *MLH1* (p.615_616del, INFORM) (**Figure 3B**).^27^ In sum, we identified 12 unique damaging germline *SMARCAL1* variants in 15 OS cases across the replication cohorts for a prevalence of 2.5% (P_Fisher_ <0.0001, OR= 7.3 [4.0 - 12.2]), which is similar to the 2.6% prevalence in the discovery cohort (**Table 1**, **Figure 3A, B; Supplementary Table S8, S9**).

Across replication cohorts, tumor WGS data were available only for the INFORM cases. We observed *SMARCAL1* LOH in all three relapsed OS tumors due to copy number variation (**Supplementary Figure S2**). Two of three tumors also had RNAseq data available, one of which (c.1335-2A>T) exhibited low *SMARCAL1* expression (**Supplementary Figure S3)**. Loss of SMARCAL1 function has been shown to result in alternative-lengthening of telomeres (ALT) in glioblastoma cells.^26^ To this end, we observed all three OS tumors in the replication cohort to be ALT positive as determined by TelomerHunter^28^ (**Supplementary Material**). In addition, we assessed tumor genomic data in these three cases for variations in ALT associated genes, including *ATRX, DAXX* and *H3F3A* and did not observe somatic or germline SNVs (n=3 WES) or reduction in RNA expression (n=2 RNAseq) in these genes. These data suggest that loss of SMARCAL1 function may induce ALT, a potential mechanism by which it promotes osteosarcoma formation (**Figure 3B, Supplementary Figure S3**).

## Discussion

Highly penetrant CPS result from germline variations in DNA damage repair (DDR) genes. To date, comprehensive studies investigating the germline landscape of DDR alterations in pediatric cancers have been lacking, in part due to the lack of case-control designs that harmonize differences such as batch effects from library preparation, sequencing platforms, and variant calling pipelines necessary for identifying novel cancer-predisposing variants. To circumvent these issues, in this study we re-mapped raw sequencing data and performed joint genotype calling across 5,993 pediatric primary cancer cases and 14,477 adult non-cancer controls. Further, we only included variants based on stringent criteria, including rarity in the general population and presumed negative impact protein expression or function based on in-silico pathogenicity predictions and prior interpretations available through established variant curation databases such as ClinVar. Through this approach, we established a 28.3% prevalence of putative damaging variants in DDR genes across a wide array of childhood cancers. Statistical testing confirmed known associations, including germline *TP53* PV in ACC and HGG, and *BARD1* PV in NBL. Importantly, we discovered novel associations of *POLL* PV in EWS, *SMC5* PV in MB, and *SMARCAL1* PV in OS. Using three independent replication cohorts, we confirmed statistical enrichment of germline damaging variants in *SMARCAL1*, highlighting it as a novel predisposing gene in which damaging variants increase the risk for pediatric OS.

We find that 2.5% of OS cases harbor germline *SMARCAL1* variants of which two-thirds are expected to cause haploinsufficiency due to protein truncation while one-third are missense and predicted to be damaging. Ballinger et al., previously reported germline truncating *SMARCAL1* variants in 19 sarcoma cases including two OS, ^29^ while Akhavanfard et al., who used data from the SJLIFE cohort, identified *SMARCAL1* in three OS cases.^30^ Moreover, two cases of OS have been reported in individuals with Schimke immune-osseous dysplasia (SIOD), a multisystemic disorder secondary to biallelic *SMARCAL1* loss-of-function alterations that is typified by bony defects (e.g., short stature, bony dysplasias) suggesting that OS is a cancer associated with *SMARCAL1* dysfunction.^31^ However, the rarity and short life span (median age of death: 11years) of individuals with SIOD preclude us from accurately assessing the prevalence and penetrance of OS in this context. No other germline alterations in OS-relevant CPG were identified in our cases, which supports *SMARCAL1* as an independent risk factor for OS.^32^ Six of 12 osteosarcoma cases (50%) with available clinical information experienced disease relapse. Longitudinal studies are needed to define the penetrance and clinical features characteristic of OS associated with germline *SMARCAL1* variation.

Somatic LOF mutations in *SMARCAL1* have been identified in glioblastoma resulting in SMARCAL1 deficiency and permitting ALT-mediated telomere synthesis, a homologous recombination-based mechanism of telomere elongation utilized by 10-15% of high-risk cancers.^26,33^ Consistent with these findings, ALT appeared active in all three tumors from INFORM with bi-allelic *SMARCAL1* alterations. ALT-positive tumors also frequently harbor LOF mutations in chromatin modification genes including, *ATRX*, *DAXX* and *H3F3A*, which contribute to ALT induction by dysregulating histone H3.3 deposition at telomeres.^34^ While we did not observe alterations in these genes in the three tumors with bi-allelic *SMARCAL1* mutations, one of two tumors with available data in our discovery cohort harbored an inactivating *ATRX* mutation. Ballinger et al., also reported LOH in 5 of 19 sarcomas with germline *SMARCAL1* variants; however, it is not possible to discern whether these were OS cases.^29^ Altogether, these data suggest that *SMARCAL1* functions as a tumor suppressor. We propose a model whereby germline *SMARCAL1* PV cause partial or altered SMARCAL1 function, leading to DNA replication and repair defects throughout the genome, including the telomeres, with acquisition of second somatic hits in ALT permissive genes, which results in OS tumor formation (**Figure 3D**).

In addition to *SMARCAL1*, we report a novel association of *SMC5* in group 3 MB and *POLL* in EWS. Out of four individuals with germline *SMC5* PV, one harbored a second somatic hit in *SMC5*, resulting in a late replication error mutational signature, which may correlate with this patient’s aggressive disease features (metastatic disease at diagnosis, relapse, death). Future studies in larger MB cohorts are required to validate this association and the role of SMC5 dysfunction in high grade MB. Limited data are available for the role of the polymerase lambda gene, *POLL,* in cancers, including EWS. Loss of *POLL* leads to BER deficiency in mouse embryonic fibroblasts.^35^ Tumors harboring the EWS driver, *EWSR1::FLI1*, which was observed in one of two *POLL* germline mutated EWS tumors, are significantly sensitive to the PARP inhibitor olaparib, which is further accentuated in the presence of other DNA damaging agents.^36^ Although clinical response to PARP inhibitors in EWS is underwhelming, cases where the BER pathway is impaired, such as those with *POLL*-mediated predisposition, may constitute a subset with improved outcomes.

The following limitations should be considered when interpreting the results of this study. First, to achieve adequate statistical power, we combined high-risk primary cancers with presumably lower risk cancers from adult survivors of childhood cancer. This approach may have diluted genetic signals in primary malignancies and added survival bias when examining the risk of childhood cancer, which warrants reassessment of primary cancers as larger cohorts become available. In our study, the variant filtering criteria were rigorous, and it is possible that additional clinically relevant germline variants with lower REVEL scores remain undescribed. To test this in our INFORM replication cohort, we decreased the filtering REVEL score to 0.5 and identified one germline *SMARCAL1* missense variant (p.D424V) in two unrelated OS cases. Matched tumor data from these cases revealed somatic second hits in the remaining wild-type *SMARCAL1* allele; however, we have not included these cases in our statistical analyses to maintain consistency in how the discovery and replication cohorts were analyzed. Future studies are warranted to determine the extent to which these and other germline *SMARCAL1* variants play a role in OS tumorigenesis. Notably, associations of *SMARCAL1* with OS and *SMC5* with MB remain statistically enriched even when analyses are restricted to include only protein-truncating and splicing variants (**Supplementary Table S5**). Finally, we could not establish the mode of inheritance or co-segregation with other cancers for many of the identified variants due lack of familial testing. Future efforts examining the relatives of germline *SMARCAL1* PV carriers are needed as this information may serve to strengthen evidence of pathogenicity. Finally, assessment of tumor genomic data is critical for determining the functional consequence of predisposing variants; however, these data were available for only a limited number of cases.

In summary, outcomes for children with OS, especially those with relapsed or metastatic disease, remain suboptimal. Our finding that germline mutations in *SMARCAL1* predispose to OS serves as a foundation for future studies aimed at developing novel therapies for this aggressive cancer, one for which there have been little advancements in treatment over the last four decades.^37^ Similarly, genetic testing for germline *SMARCAL1* PV will enable prospective surveillance for the detection and treatment of incipient OS tumors at their earliest and most curable stages.

## Acknowledgements

Funding for this study was provided by the American Lebanese Syrian associated charities. This study was supported by the following National Cancer Institute grants, R01CA283333 (Zhaoming Wang and Kim E Nichols), The St. Jude Lifetime Cohort (SJLIFE) (CA195547, M.M. Hudson, K.K. Ness), and The Childhood Cancer Survivor Study (CCSS) (CA55727, G.T. Armstrong). The INFORM program is financially supported by the German Cancer Research Center (DKFZ), several German health insurance companies, the German Cancer Consortium (DKTK), the German Federal Ministry of Education and Research (BMBF), the German Federal Ministry of Health (BMG), the Ministry of Science, Research and the Arts of the State of Baden-Württemberg (MWK BW); the German Cancer Aid (DKH), the German Childhood Cancer Foundation (DKS), RTL television, the aid organization BILD hilft e.V. (Ein Herz für Kinder) and the generous private donation of the Scheu family. We would like to express our sincere thanks to Carsten Maus, Erjia Wang (Next Generation Sequencing Core Facility, DKFZ). Lena Weiser, Gregor Warsow (Omics IT and Data Management Core Facility, DKFZ) for their highly dedicated support in data management and processing and Rolf Kabbe (Division of Pediatric Neurooncology, DKFZ) for his sincere and dedicated contribution to the bioinformatics analyses. Biostatistics support is provided by the Biostatistics Shared Resource (BSR) of the St. Jude Children’s Research Hospital and St. Jude Comprehensive Cancer Center (NIH P30CA021765). The authors thank the patients and families included in this study and members of the St. Jude Clinical Genomics Laboratory, without whom this work would not have been possible.

## Data Availability Statement

The processed genomic data generated in this study are provided in the **Supplementary Tables**. Controlled access raw genomic data can be requested via St. Jude Cloud at https://platform.stjude.cloud/. The Childhood Cancer Survivor Study is a US National Cancer Institute funded resource (U24 CA55727) to promote and facilitate research among long-term survivors of cancer diagnosed during childhood and adolescence. CCSS data are publicly available on the St Jude Survivorship Portal within the St. Jude Cloud at https://survivorship.stjude.cloud/. In addition, use of the CCSS data that leverages the expertise of CCSS Statistical and Survivorship research and resources will be considered on a case-by case basis. For this use, a research Application of Intent followed by an Analysis Concept Proposal must be submitted for evaluation by the CCSS Publications Committee. Users interested accessing this resource are encouraged to visit http://ccss.stjude.org. Full analytical data sets associated with CCSS publications since January 2023 are available on the St. Jude Survivorship Portal at https://viz.stjude.cloud/community/cancer-survivorship-community~4/publications. Any additional data are available upon request from the corresponding author.

## Notes

Conflict of Interest: No author disclosures were reported.

### Competing Interest Statement

The authors have declared no competing interest.

### Funding Statement

Funding was provided by the American Lebanese Syrian Associated Charities and US National Institutes of Health, German Cancer Research Center, German Cancer Consortium.

### Author Declarations

IRB of St. Jude Children's Research Hospital gave ethical approval for this work.

### Summary of Updates

Removed sections from abstract that did not belong in the abstract format. Other minor edits to figures.

