## Supplementary Figure for "Comprehensive investigation of DNA damage repair genes in children with cancer identifies *SMARCAL1* as novel osteosarcoma predisposition gene"

### Supplementary Material

#### Supplementary Methods

##### Clinical metadata curation and data harmonization

Clinical metadata for RTCG, G4K, SJLIFE, PCGP cohorts was downloaded from the St. Jude Cloud genomics platform (<https://platform.stjude.cloud/data/diseases/paired-tumor-normal>) and for TARGET cohort, metadata was downloaded from the NCI-GDC portal (<https://portal.gdc.cancer.gov/projects/>). Cancer type annotations were reviewed by a team of physicians, clinical research associates, and bioinformatics scientists to collapse down to 22 principal cancer types (level 2) and 3 cancer classes (hematologic, brain, solid tumors).

##### Variant Calling and Filtering

Raw sequencing reads were obtained from St. Jude’s data repository (RTCG, G4K, SJLIFE, PCGP) and dbGap (TARGET, 1000 genomes, ADSP). All samples were uniformly mapped to the GRCh37-lite reference genome using BWA^1^. GATK best practices workflow was used to perform variant calling (HaplotypeCaller) and joint genotyping (GenotypeGVCFs).

We retained high quality variants that passed filtering using following criteria: allelic balance >0.2, genotype quality >20, variant allele frequency (VAF) for heterozygous variants between 20-80%, minimum of 10 alternate reads supporting single nucleotide variants (SNVs) and 7 alternate reads supporting InDels, and missingness <25% of samples. We performed variant annotation using ANNOtate VARiation (ANNOVAR)^2^. We also annotated all the variants using InterVar^3^ automated clinical interpretation based on the American College of Medical Genetics and Genomics (ACMG) guidelines.^4^ We first filtered for rare variants (gnomAD v2.1^5^, noncancer subset allele frequency <0.05%). Genetic ancestry and sex of all samples was determined by extracting 100 principal components from this data and aligning it with major populations from the 1000 genomes project.

##### Tumor molecular characterization in discovery cohorts

We used clinical genomics data (RTCG, G4K) and published catalogs of P/LP variants (PCGP, TARGET, SJLIFE, G4K) to determine additional somatic mutations for the 13 cases where matched tumor WES data were available. We calculated the expression quantiles for each gene using normalized expression of all cases within a cancer type and ‘ecdf’ function (R package stats v3.6.2). The Sashimi plot for SJMB030089 and an unrelated MB case without *SMC5* alteration used as ‘control’ was generated using Integrative Genomics Viewer (IGV).^6^

For somatic mutational signature analysis, we performed somatic mutation calling using Mutect2^7^, SomaticSniper^8^, VarScan2^9^, Strelka2^10^, and MuSE^11^ followed by consensus calling with at least 2 callers. We used sigProfiler^12^ package to extract mutational signatures from the five samples. We used signature.tools.db^13,14^ package to run 1,000 permutations on the original 96-element mutational catalogue of each sample and estimate the signature activities in each permutation. Signatures with relative contribution above 5% with a *P* value <0.01 were considered significant and other signatures were annotated as “Unassigned”. The final signature activities of all 21 samples were summarized into a plot using R package ComplexHeatmap.^15^

##### Replication cohort sequence analysis

Whole Exome Sequencing (WES) data from INFORM and GCCR were processed using an in-house DKFZ workflow. Matched tumor and control samples were aligned to the human reference genome (build 37, version hs37d5) using the DKFZ One Touch Pipeline (OTP).^16,17^ Single-nucleotide variants (SNVs) were detected using the SNVCalling Workflow (version 1.2.166-3), which is based on samtools^18^ and bcftools^19^, incorporating parameter modifications and heuristic filtering as described.^20^ Small insertions and deletions (InDels) were identified with the IndelCalling Workflow (version 2.4.1), which is based on Platypus.^21^ Variant call confidence was assessed using Platypus, considering only variants with a confidence score between 8 and 10. Functional annotation of SNVs and InDels was performed using ANNOVAR.^2^ INFORM neuroblastomas and Ewing Sarcomas were called using freebayes v1.1.0. Copy number variations (CNVs) were identified using CNVkit (version 0.9.3)^22^ with default parameters. B-allele frequency output files from CNVkit were used to generate custom BAF plots. Additionally, concurrent low-coverage whole-genome sequencing (lcWGS) or deep-coverage whole-genome sequencing (WGS) samples were analyzed using qDNAseq and the resulting IGV formatted segments file was used to generate custom copy number plots.

RNA sequencing data were processed through the DKFZ OTP RNA sequencing workflow (version 1.3.0).^17^ Read quality was assessed using FastQC, and sequences were aligned to the reference genome (hs37d5) with the STAR aligner (version 2.5.3a).^23^ Gene-level quantification was performed using featureCounts^24^, considering only exon regions defined in GENCODE v19 and including both reads from paired fragments.

ALT status of low coverage whole genome sequencing data (lcWGS) was assessed using TelomereHunter^25^, without applying a strict threshold. Samples were classified as ALT-positive if the tumor exhibited a higher telomere count than the matched normal along with enrichment of variant repeats above the expected log2 ratio. Cases with only one of these two metrics positive were considered borderline, as this pattern can occur in tumors with TERT activity or very low tumor purity. Samples negative for both metrics were classified as ALT-negative. Additionally, if a sample was positive for TERT overexpression this was used as an additional criterion to determine ALT-positivity.

##### Statistical analysis

We performed QQ-plot analysis using both the selected list of 189 DDR genes and a randomly selected list of 1000 genes for both Firth and GLM regression analysis (**Supplementary Figures S4, S5**). We report associations for cancer types that do not show significant deviations from normality in QQ-analysis. To compare the distribution of ages at diagnosis between cases with or without germline damaging DDR gene variants in specific cancer types, we used the Wilcoxon rank-sum test as the normality assumption was not met.

#### Supplementary Figures

##### Supplementary Figure S1: Molecular features in carriers of predisposing variants

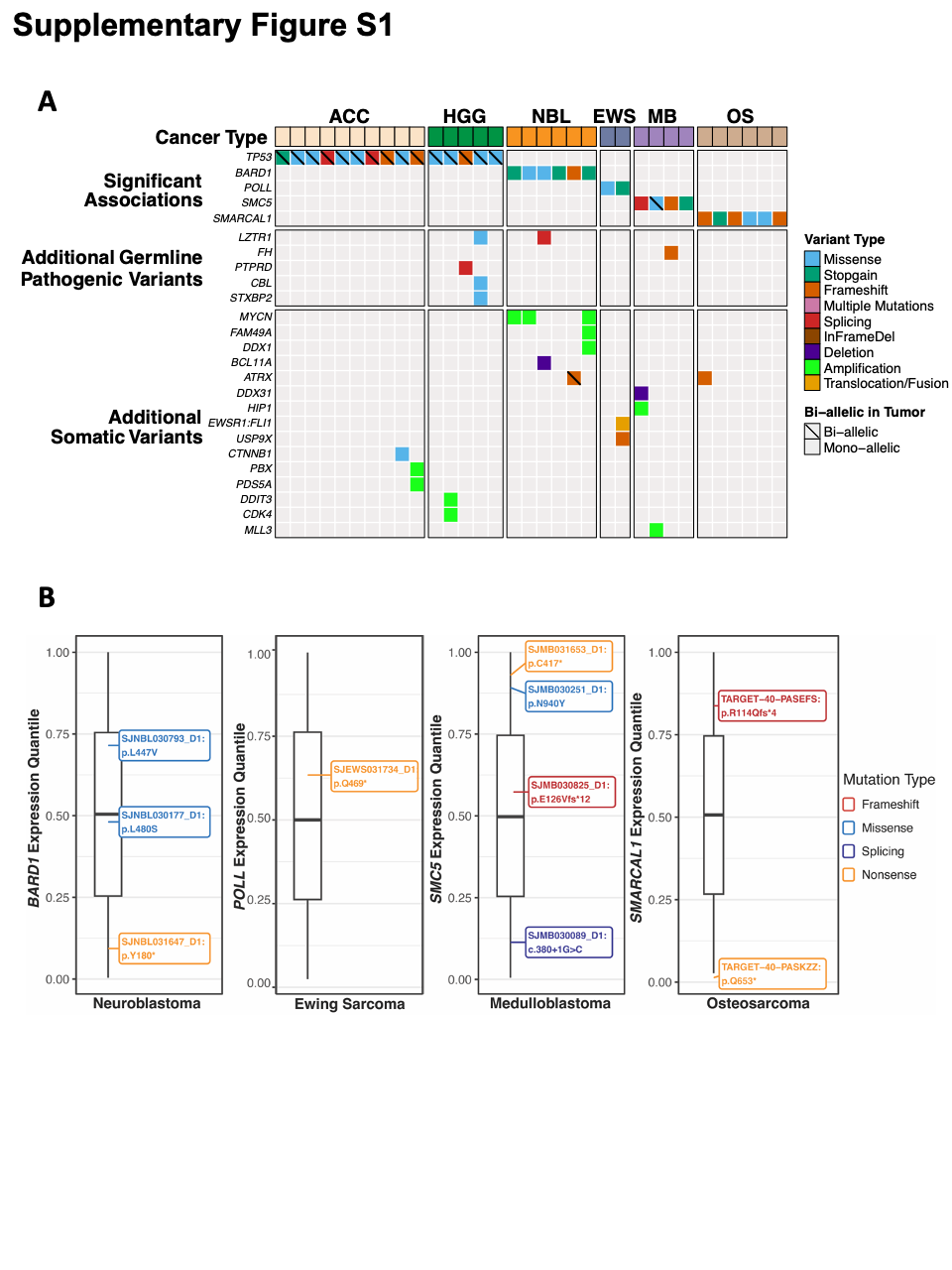

(**A**) Mutational landscape of TP53, BARD1, POLL, SMC5, and SMARCAL1 across adrenocortical carcinoma (ACC), high-grade glioma (HGG), neuroblastoma (NBL), osteosarcoma (OS), medulloblastoma (MB), and Ewing sarcoma (EWS). The heatmap displays significant genetic associations, additional germline pathogenic variants, and additional somatic variants identified in each cancer type. Variant types include missense (blue), stop-gain (red), frameshift (orange), multiple mutations (pink), splicing (purple), deletion (green), amplification (yellow), and translocation/fusion (brown). Tumor samples with biallelic mutations are indicated by diagonal lines, while monoallelic variants remain unmarked.

(**B**) Expression quantile distribution of BARD1, POLL, SMC5 and SMARCAL1 mutations across tumor types. Boxplots correspond to the normalized RNA expression quantiles calculated using ‘ecdf’ function in R for all RNAseq RSEM values within each cancer type. Different mutation types are shown: frameshift (red), missense (blue), splicing (purple), and nonsense (orange). Data is presented for neuroblastoma (BARD1), osteosarcoma (SMARCAL1), and medulloblastoma (SMC5)

##### Supplementary Figure S2: Tumor copy number and B-allele frequency plots for three relapsed tumors from INFORM cohort

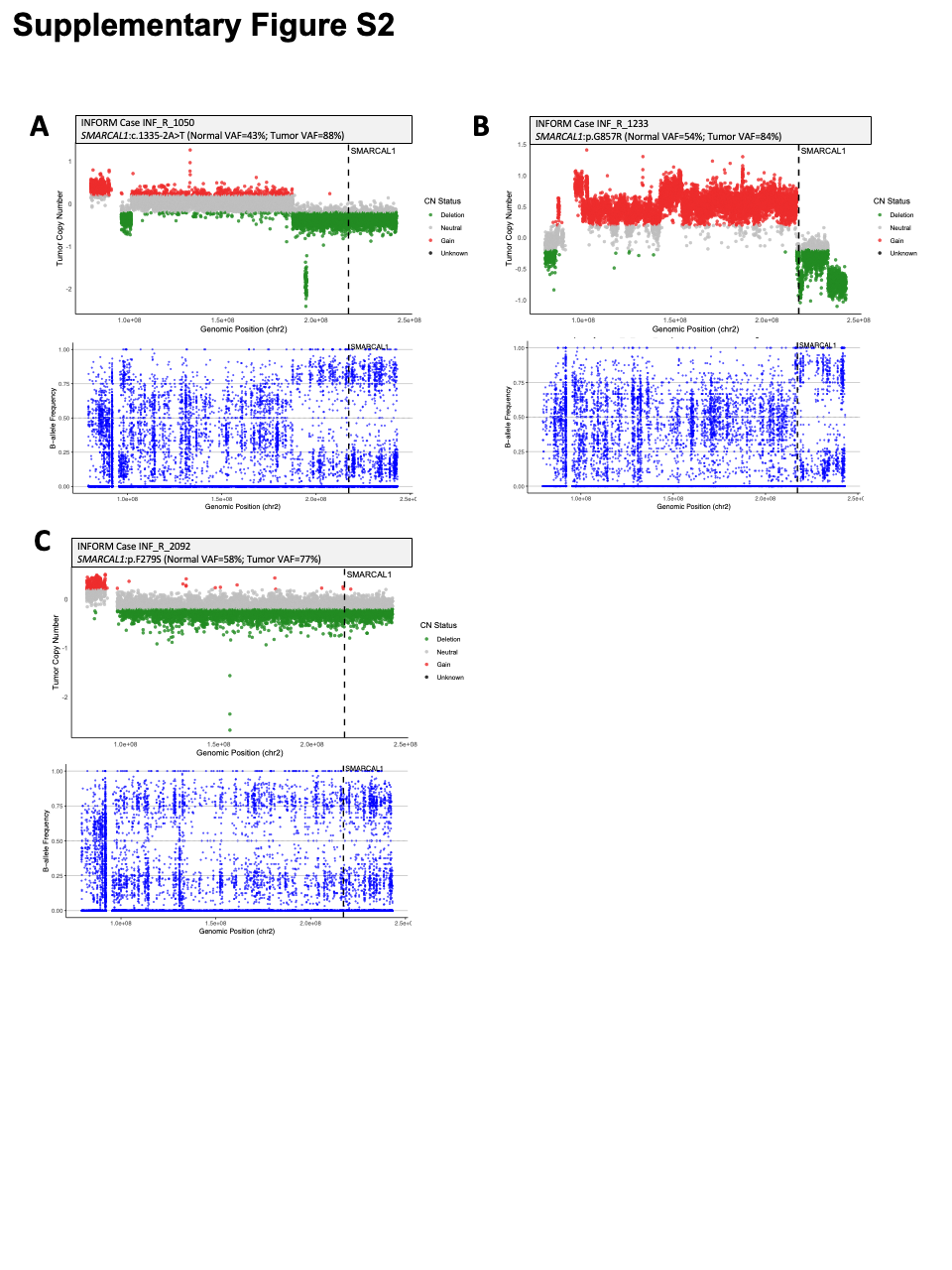

(**A-C**) For each of the three cases from INFORM cohort, tumor WGS data was analyzed. Top panel shows tumor copy number at the SMARCAL1 locus (dashed line) indicated as copy number gain (red), deletion (green), neutral (gray), or unknown (black) CN status. Respective germline variants in SMARCAL1 are shown along with corresponding tumor and normal variant allele fractions (VAF). Lower panel shows corresponding B-allele frequency plots for the three samples.

##### Supplementary Figure S3: Tumor RNA expression of *SMARCAL1*, *ATRX*, *DAXX*, *H3F3A*

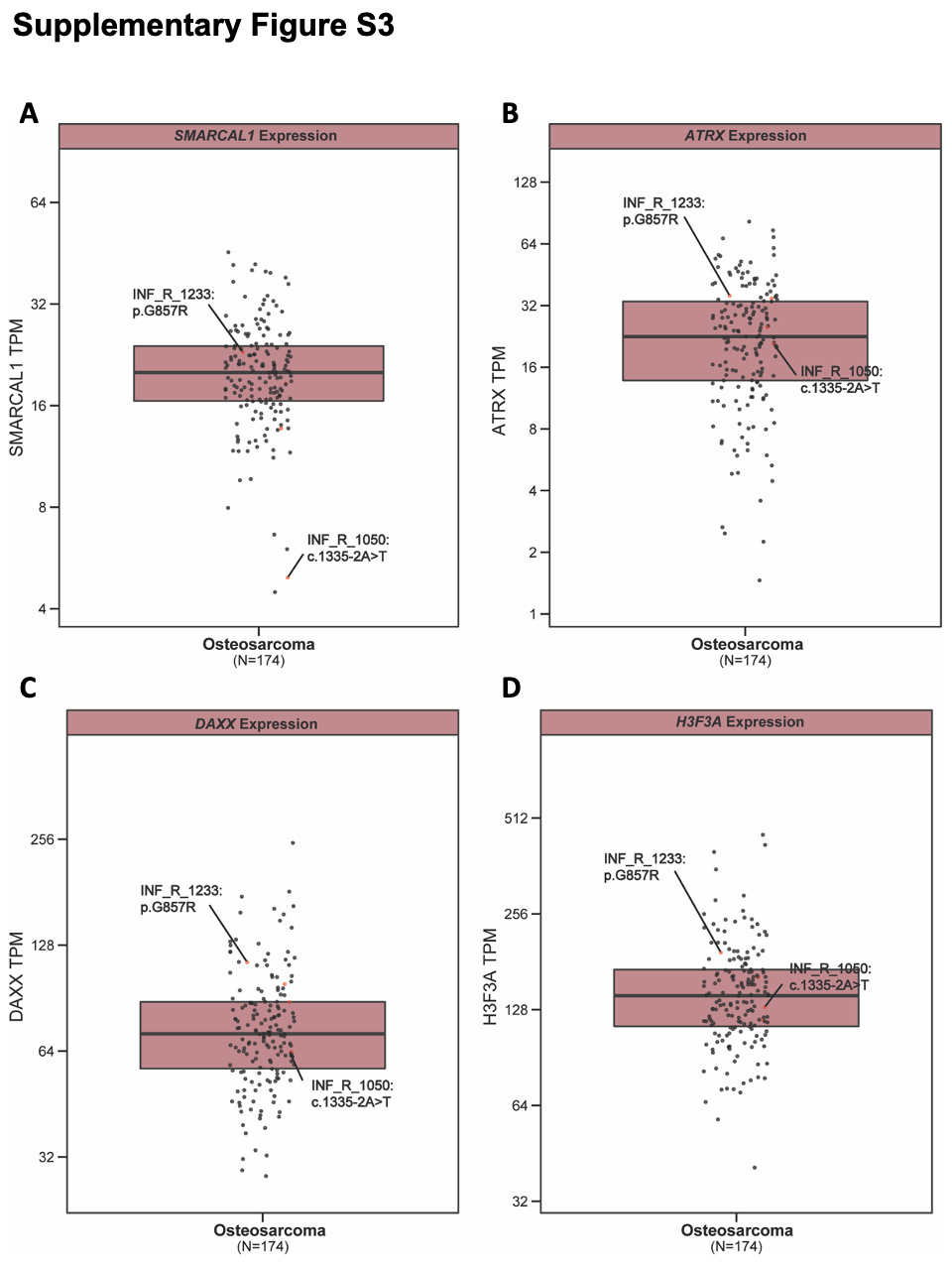

RNA expression of *SMARCAL1* (**A**), *ATRX* (**B**), *DAXX* (**C**), *H3F3A* (**D**) in OS tumors from INFORM. Two cases with a germline damaging *SMARCAL1* variant and somatic *SMARCAL1* LOH are labelled.

##### Supplementary Figure S4: Q-Q analysis for Firth regression using chi-squared tests (Discovery Cohort)

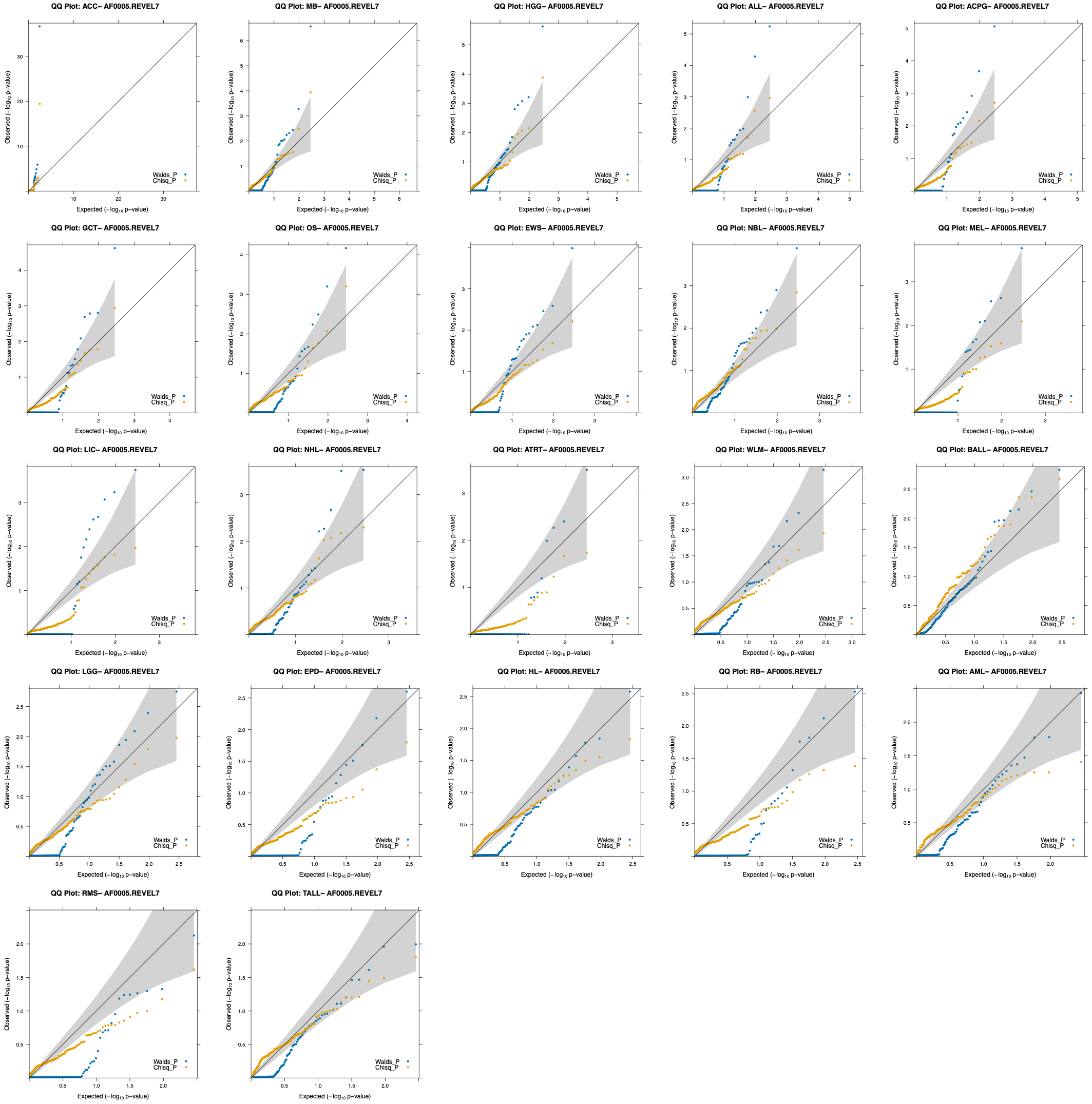

##### Supplementary Figure S5: Q-Q analysis for GLM regression tested using Walds or Chi-squared tests (Discovery Cohort)

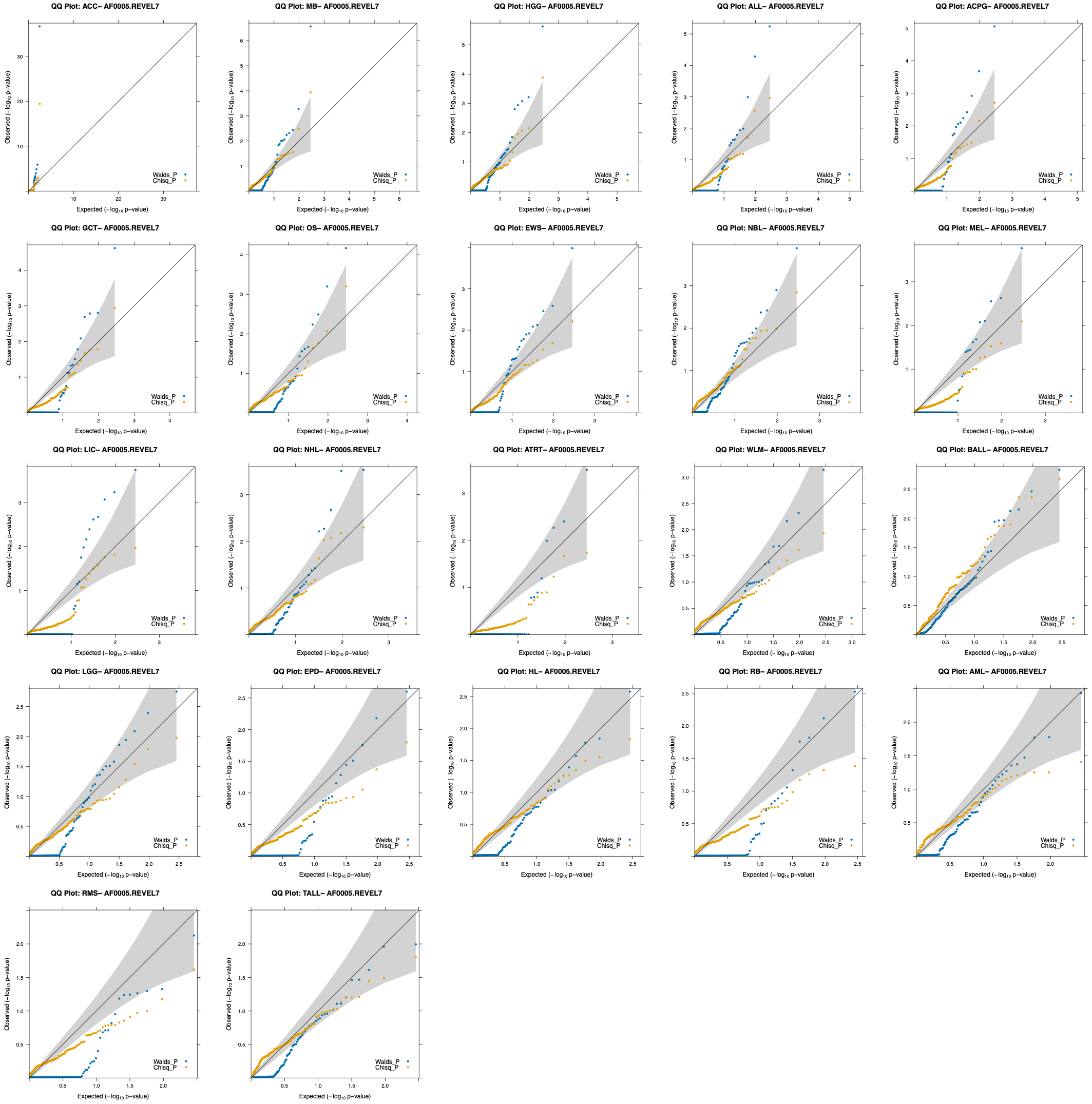

#### Supplementary Tables

All supplementary tables are provided in a separate Excel file.

##### Supplementary Table S1: List of 5,993 pediatric cancer samples in the discovery cohort

##### Supplementary Table S2: List of six DDR pathways and 189 unique genes analyzed in this study

##### Supplementary Table S3: Ancestry composition of 22 pediatric cancers and controls

##### Supplementary Table S4: List of 2,059 pathogenic variants in DDR genes across all cancers

##### Supplementary Table S5: Rare variant association analysis summary

##### Supplementary Table S6: Predisposing variants in significant DDR genes in the discovery cohort

##### Supplementary Table S7: Age of cancer onset in carriers and non-carriers of DDR variants

##### Supplementary Table S8: Rare variant enrichment analysis in replication cohorts

##### Supplementary Table S9: Summary of germline *SMARCAL1* variants across all cohorts
